# Predictors of low birth weight in pregnant women with malaria: a prospective cohort facility-based study in Webuye-Kenya

**DOI:** 10.1101/2023.10.09.23296556

**Authors:** Joseph Mukala, Dominic Mogere, Peter Kirira, Bernard Kanoi, Violet Akinyi, Francis Kobia, Harrison Waweru, Jesse Gitaka

**Author notes:** **Correspondence** (Joseph Mukala, MD, MPH, P.O. Box 342-01000, Thika, Kenya. Cellphone).

## Abstract

Malaria is caused by protozoa of the genus *Plasmodium* and remains a major public health burden in Sub-Saharan Africa. Its prevalence varies between 9 to 18% with harmful consequences to both the mother and her baby, including adverse pregnancy outcomes such low birth weight, high morbidity, and mortality. However, effective antenatal strategies for improving maternal and child health outcomes through the prevention, early detection, and treatment of malaria in pregnancy, are still lacking in resource-constrained settings. Here, we sought to determine the predictors of low birth weight in pregnant women with malaria in a cohort study in Webuye hospital. Prior to the enrollment of 140 participants, permission was sought from relevant institutions and consent from the participants. Malaria test was conducted either with microscopy or rapid test, and then the cohort splits into malaria positive and negative followed up from the first antenatal visit (March 2022) and delivery (December 2022). Before data collection, training, pre-testing and quality control were duly observed. Data were fed into SPSS 27 version, Chi-square and Fischer’s Exact were used for bi-variate analysis at a p-value less or equal 0.05 (95%). Our results revealed that birth cohort with malaria did not result in significant low birth weight with a relative risk of 0.999, confidence level of 0.926-1.077. The prevalence of low birth weight was 4.6% with 6 cases of which 3 (4.5%) in the negative cohort and 3 (4.7%) in the positive cohort. Anemic pregnant women were 41 (31.5%), HIV were 5 (3,8%), pre-eclampsia was 5 (3.8%), gestational diabetes was 2 (1.5%). Confounding factors, such as anemia, HIV, preeclampsia, and gestational diabetes did not influence low birthweight (p-value >0.923). Otherwise, most of the participants were aged 18–25 years, were primigravida, were married, had secondary school level education, earned between 20-30 thousand shillings, were resident in rural areas, and were in their second trimester. Marital status, gestational age and area of residence were associated with malaria with a p-value less than 0.001 and 0.028 respectively.

## Introduction

Malaria infects approximately 515 million people in Latin America, Asia and Sub-Saharan Africa region with one to three million deaths each year [1]. Recently, malaria has affected 228 million people worldwide, with approximately 213 million in sub-Saharan Africa representing 93% of the total population [2]. Statistics show that 9.6 million people representing 19% were at risk of malaria in 2019 in the high altitude or highland zones. Bungoma, Kakamega and Baringo are in this region, and they are also endemic zones along Lake Victoria. The coastal region had 13.7 million representing 27% of the population at risk of malaria. The seasonal malaria transmission zone is in northern and central Kenya; the number of exposed persons were 11 million representing 23% of the population at risk of malaria. Nairobi and its environs have 15 million of population representing 35% were at low risk [3]. The most vulnerable persons being children and expectant women with consequences ranging from deadly complications such as anemia, abortion, intrauterine fetal retardation, small gestational for age, prematurity and low birth weight. Indeed, the last decade was characterized by the effort to reduce malaria incidence from 71 to 57 over 1000 cases in the high risk zones [4]. Worldwide, low birth weight represents 15% to 20 % of the disease burden. South-Asia and Sub-Saharan Africa are the most hit with 96% of cases of malaria [5]. Among as many as eleven million pregnant women who were exposed to malaria infection in 2018, the consequences translated to an estimated 872,000 low birth weights newborns being the highest record of 16% in the Western Africa region compared to the central and Eastern Africa [6]. Interestingly, countries that recorded less than hundred cases of malaria among autochthones population increased from seventeen in 2010, to twenty-five in 2017, and finally twenty-seven in 2018. Moreover, Algeria and Malaysia have not so far reported cases of malaria while China has been awarded a certificate of malaria elimination by WHO [7], [8]. The Kenya National Malaria Program working closely in partnership with other supportive agencies to assist the districts and counties as the execution level of ensuring the smooth process in line with prevention, detection, management of malaria cases based on WHO recommendations [3]. In another research study carried out in Kenya in 2015 on socio-demographic factors highlighted determinants to understanding the reason why malaria is still a big challenge and public health threat in rural areas than in the urban areas. The study found that rural areas are associated with long bushes, stagnant water along houses, which provide good breeding ground for mosquitoes [10]. Malaria physiopathology begins with sequestration of merozoite/sporozoite in the placenta, with consequences reduced placental perfusion and release of free radicals such as soluble endoglin, cytokines, soluble kinase tyrosine leading to fetal growth restriction, stillbirth, low birth weight, prematurity which will be complicated with high morbidity and mortality. This study suggested that biomarkers to identify placental suffering may be combined to increase their specificity [11]. Pregnant women and malaria study carried out in Bungoma County found a prevalence of 21.6% with high likelihood of infection during the first trimester of gestation when compared with other trimesters with *Plasmodium* falciparum representing 83%, the density of the parasite in the thin blood film was higher for the same type. The study recommended continuous screening, case management and preventive measures, as well as continuous health education session to curb malaria infection [12].

## Materials and methods

### Study design

The study design was a prospective cohort conducted at Webuye hospital from (March 2022) to (December 2022), either 10 months. Participants were enrolled from 16 weeks of pregnancy and followed up to the delivery.

### Study setting

The study was conducted in Bungoma County Code 39, Sub-County Webuye West code 3911, Webuye hospital. The County has a population estimated at 1,919,490 with 939,105 males and 980,385 females, 429,762 women of childbearing aged between 15-49 years. There are 12 sub-Counties, 45 wards and 149 Sub-locations. The County covers an area of 3032 km^2^ and lies between latitude 00 28’and latitude 10 30’ North of the Equator, and longitudinal 340 20’East and 350 15’East of the Greenwich meridian. It borders the Republic of Uganda to the Northwest, Trans-Nzoia County to North-East, Kakamega County to the East and South-East, and Busia County to the West and Southwest. It is characterized by two rainy seasons, a long rainy season goes from March to July, and a short season from August to October with an annual rainfall ranging between 400 mm to 1,800 mm. The temperature varies between 0^0^C and 320^0^C [13], [14].

### Sampling frame and inclusion criteria

In this study, a total of 140 pregnant women aged between 18-49 years with gestation at 16 weeks were selected from ANC. To be eligible for the study, participants needed to be mentally stable and residents in the area for almost six months. Malaria testing was conducted using either microscopy or rapid diagnostic test. Out of the 140 participants 70 (50%) tested positive for malaria while the other tested negative.

### Data collection

Data were collected using the following steps: The questionnaire was pretested and administered to participants in English, with Kiswahili translations provided for those with language barriers. Questions were appropriately formulated, numbered, and provided with options for both close-open and open-ended answers. Socio-demographics variables were: age groups (18-25, 26-33, 34-41, 42-49), gestation (first:1-12, second:13-25, third:26-38 weeks), education level (none, primary, secondary, college/university), income earning (low:10-20Ksh, middle:21-35Ksh, high: >35Ksh), marital status (married, divorced, single, widowed, monogamous, polygamous), residence (rural, urban), occupation (housewife, employed, self-employed), residence (rural, urban), distance to the health facility (in hour or minutes). The outcome variables: low birth weight (<2500 g), normal birth weight (>2500), normal delivery (alive newborn). The following registers were used: MOH ANC (405), Maternity register (333), laboratory register (204).

### Detection and quantification of malaria parasites Microscopy

The blood sample collected from the finger prick was fixed and turned on a slide fixed with methanol 3%, and Giemsa staining within 30 minutes. Slides were examined microscopically using 100 magnifications, trophozoites/schizonts were identified over a parasitemia density measured in one microliter of blood which was calculated through quantification of malaria parasites number versus 200 WBC multiplied to 8000 to determine parasitemia one blood microliter. After 100 high-power fields visualization the results were either negative if no parasites found or positive if malaria parasites seen. Therefore, quality control and safety measures were achieved through slide re-reading by a second lab technologist, as well as infection prevention control observance.

### Rapid diagnostic test

It is immunochromatographic test to detect specific parasite antigens. The histidine-rich protein 2 (HRP2) is specific for *Plasmodium* falciparum. There is lactate dehydrogenase (LDH) or aldolase having the ability to differentiate between *Plasmodium* falciparum and non-*Plasmodium falciparum* such *ovale, vivax* and *malariae*. According to the national guideline the use of rapid diagnostic test is not recommended for follow up and cannot determine the density of parasite.

### Ethical considerations

Before enrollment participants were informed about consent process based on aspects of rights, respects, benefits, confidentiality, withdrawal and voluntarily participation. Malaria test using either microscopy or rapid test was conducted among recruited pregnant women. Ethical approval was sought from the Ethics Review Committee of Mount Kenya University, and a research permit obtained from NACOSTI (MKU/ERC/2100, license No. NACOSTI/P/22/16233), as well as local authorizations from County and Webuye hospital management.

### Sample calculation and data analysis

The sample size calculation formula for cohort was used based on the prevalence of malaria in the non-exposed group, which was estimated at 28% according to the study of Nyamu [15]. The prevalence of malaria in the exposed group was estimated at 6.1% according to the DHIS2 [13]. Beta (10%), Alpha (5%), Confidence level of 95%, Z alpha (1.96), Z beta value (1.28), Sample size for group 1 (n1=60), Sample size for each group (n1=60), Sample size for both group (n1+n2=120), Attrition (=20%), Total sample size with attrition=144.

Afterward, data were fed cleaned, interpreted, edited and coded into SPSS 27 version, Chi-square and Fisher’s Exact were computed for categorical data, and relative risk was used for low birth weight which was the outcome of interest. The confidence level Alpha or error term used in this study was 0.05 (95%).

### Socio-demographic characteristics of pregnant women

There was strict observance in provision of consent by the participants, and malaria test done after taking blood samples. The results were obtained by conducting either rapid test or microscopy in order to confirm the diagnosis. A total of 140 (100%) were tested for malaria of which 70 (50%) tested positive for malaria while others tested negative. Overall, the majority were in the 18-25 years age-group, primigravida, married, with secondary level of education, middle income level, self-employed, residents of rural areas and in the second trimester. There was significant association between marital status (p-value <0.001), gestation in weeks (p-value <0.001) and area of residence (p-value < 0.028). [Table 1].

**Table 1:**
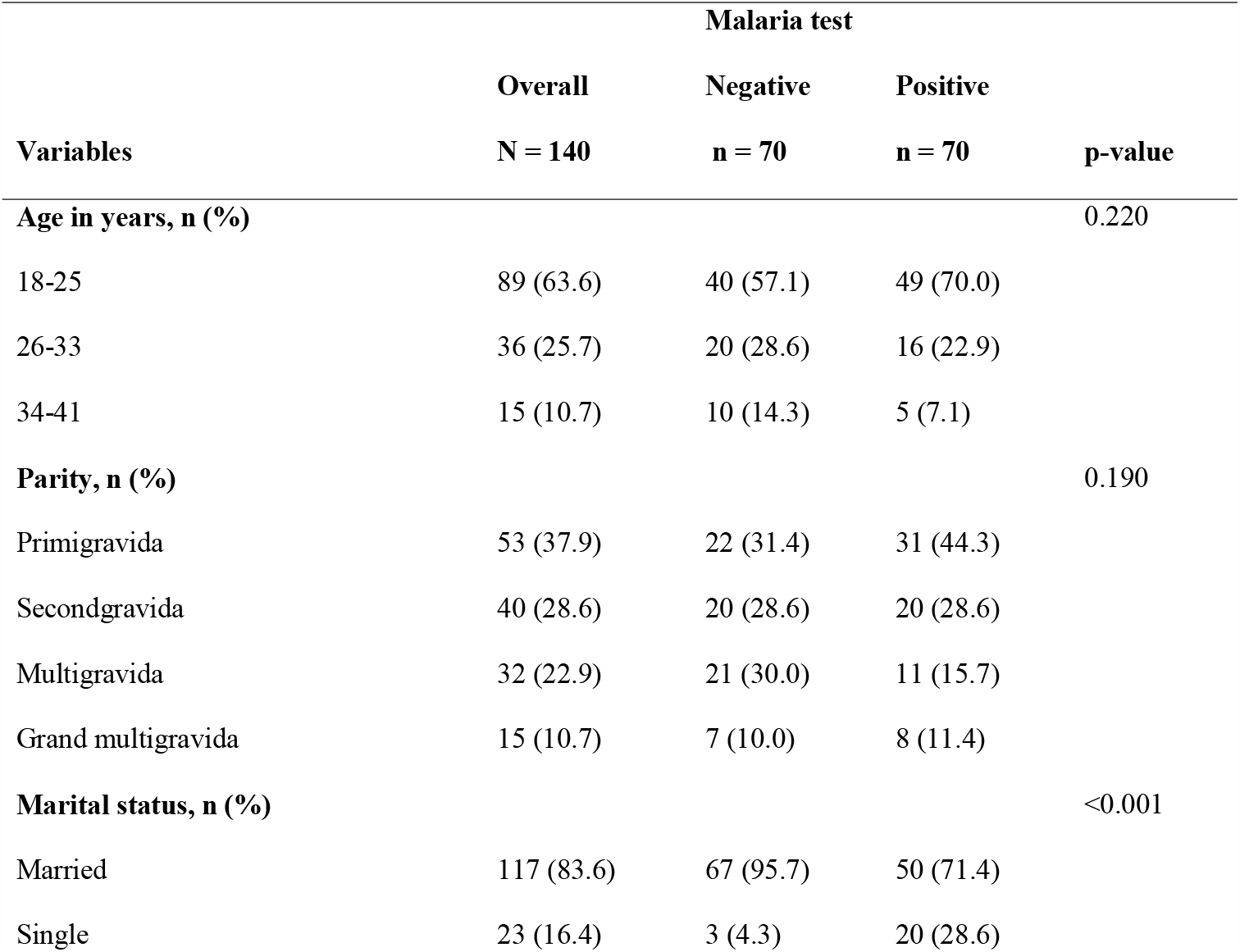
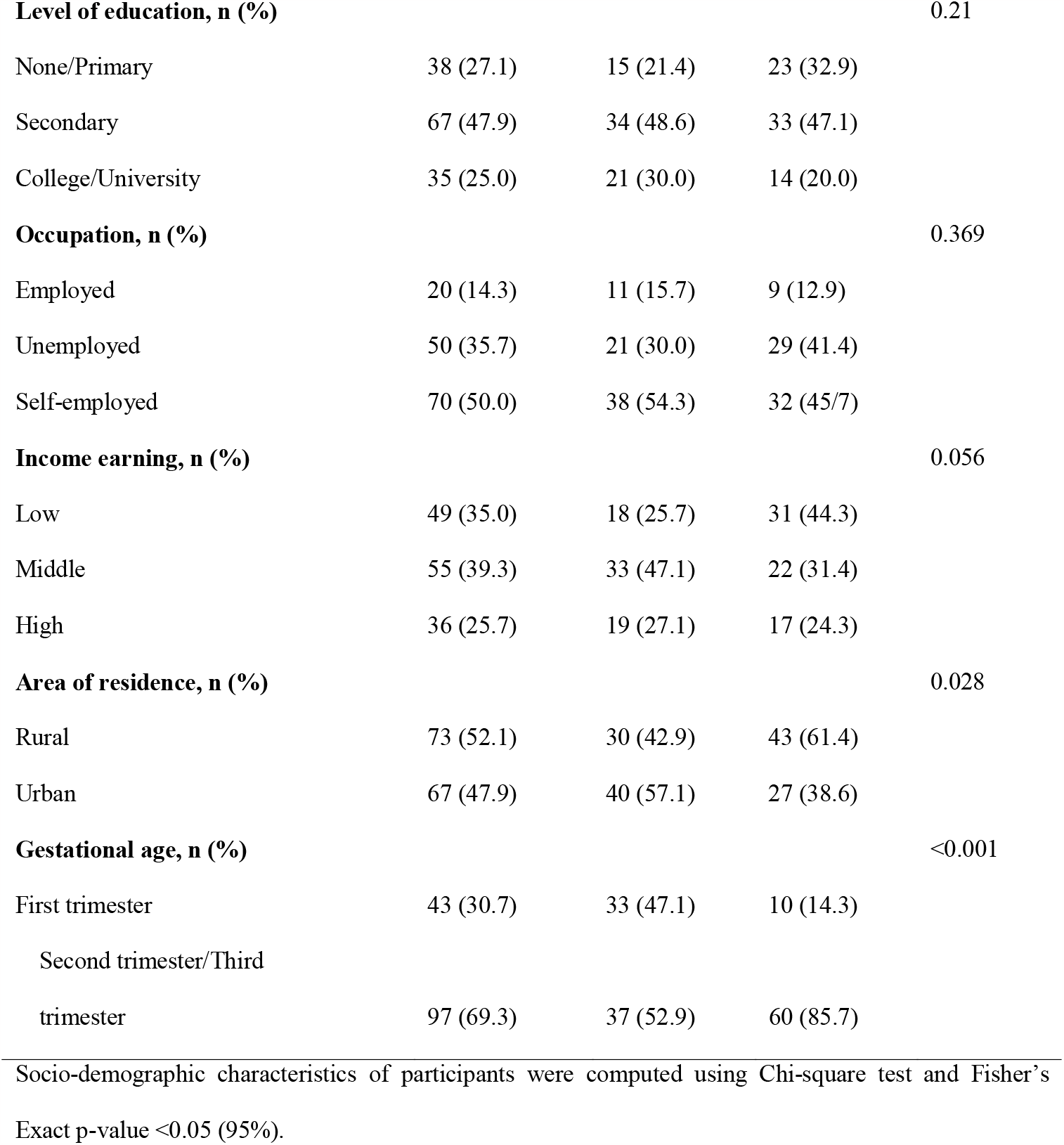
Socio-demographic Characteristics of Pregnant Women.

### Characteristics of newborns

The prevalence of low birth weight was 4.6% (6 cases). There were 140 pregnancies of whom 129 (92.2%) were live births, only 4 (2.29) were admitted, 1 (0.7%) stillbirth in the cohort of positive malaria, 10 (7.1%) were miscarriages. Very low birth weight represented 1case (0.8%) in positive malaria pregnant women and low birth weight had 5 (3.8%) and only 3 of them representing 4.7% were observed in positive malaria cohort and 2 (3.0%) in negative malaria cohort. Normal delivery with 115 (88.5%), caesarian section 15 (11.5%). Females 78 (60%) against male newborns 52 (40%). There was no statistical difference noted between the two groups. Therefore, newborns characteristics were not statistically significant in both positive and negative cohorts. [Table 2]

**Table 2:** Characteristics of Newborns.

Malaria test
| Variables | Overall, N = 140 | Negative, n = 70 | Positive, n = 70 | p-value |
| --- | --- | --- | --- | --- |

| Malaria test |  |  |  |  |
| --- | --- | --- | --- | --- |
| Variables | Overall, N = 140 | Negative, n = 70 | Positive, n = 70 | p-value |
| **Conception product outcome, n (%)** |  |  |  | 0.820 |
| Miscarriage | 10 (7.1) | 4 (5.7) | 6 (8.6) |  |
| Stillbirth | 1 (0.70) | 0 (0.00) | 1 (1.4) |  |
| Alive | 125 (89.3) | 64 (91.4) | 61 (87.1) |  |
| Admitted | 4 (2.9) | 2 (2.9) | 2 (2.9) |  |
| **Birthweight for single baby, n (%)** |  |  |  | 0.790 |
| Very low | 1 (0.8) | 1 (1.5) | 0 (.00) |  |
| Low | 5 (3.8) | 2 (3.0) | 3 (4.7) |  |
| Normal | 119 (91.5) | 61 (92.4) | 58 (90.6) |  |
| Macrosomia | 5 (3.8) | 2 (3.0) | 3 (4.7) |  |
| **Mode of delivery, n (%)** |  |  |  | 0.450 |
| Normal | 115 (88.5) | 57 (86.4) | 58 (90.6) |  |
| CS | 15 (11.5) | 9 (13.6) | 6 (9.4) |  |
| **Sex for single child, n (%)** |  |  |  | 0.830 |
| Males | 52 (40.0) | 27 (40.9) | 25 (39.1) |  |
| Females | 78 (60.0) | 39 (59.1) | 39 (60.9) |  |

Malaria diagnosis was done using either microscopy or rapid diagnosis test.

### Associated conditions versus birth weight among pregnant women

Overall, most newborns who had normal birth weight were from non-anemic women 89 (68.5%, 95% CI: 0.917 – 1.081), were non-reactive 125 (96.2), normotensive 125 (96.2) and non-diabetic 128 (98.5). HIV was diagnosed in five mothers giving a prevalence rate of 3.8%. None of enumerated conditions including malaria test were not significantly associated with birth weight (Fisher’s exact with p-value > 0.9) and Relative Risk = 0.996 at 95% C.I:917-1.081. [Table 3]

**Table 3:**
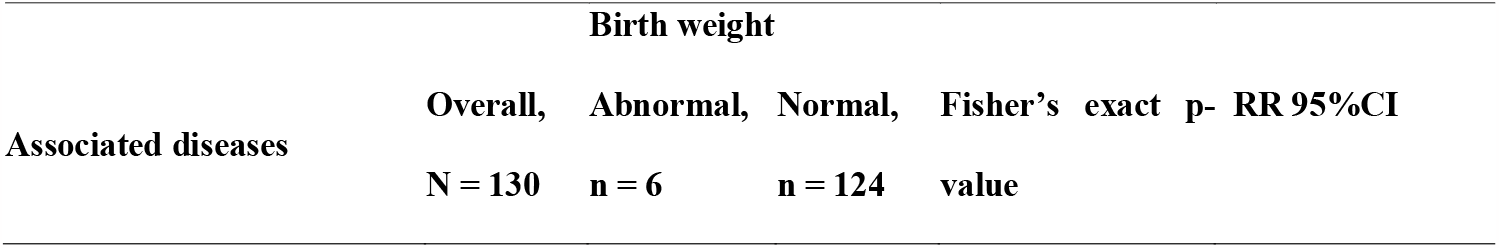
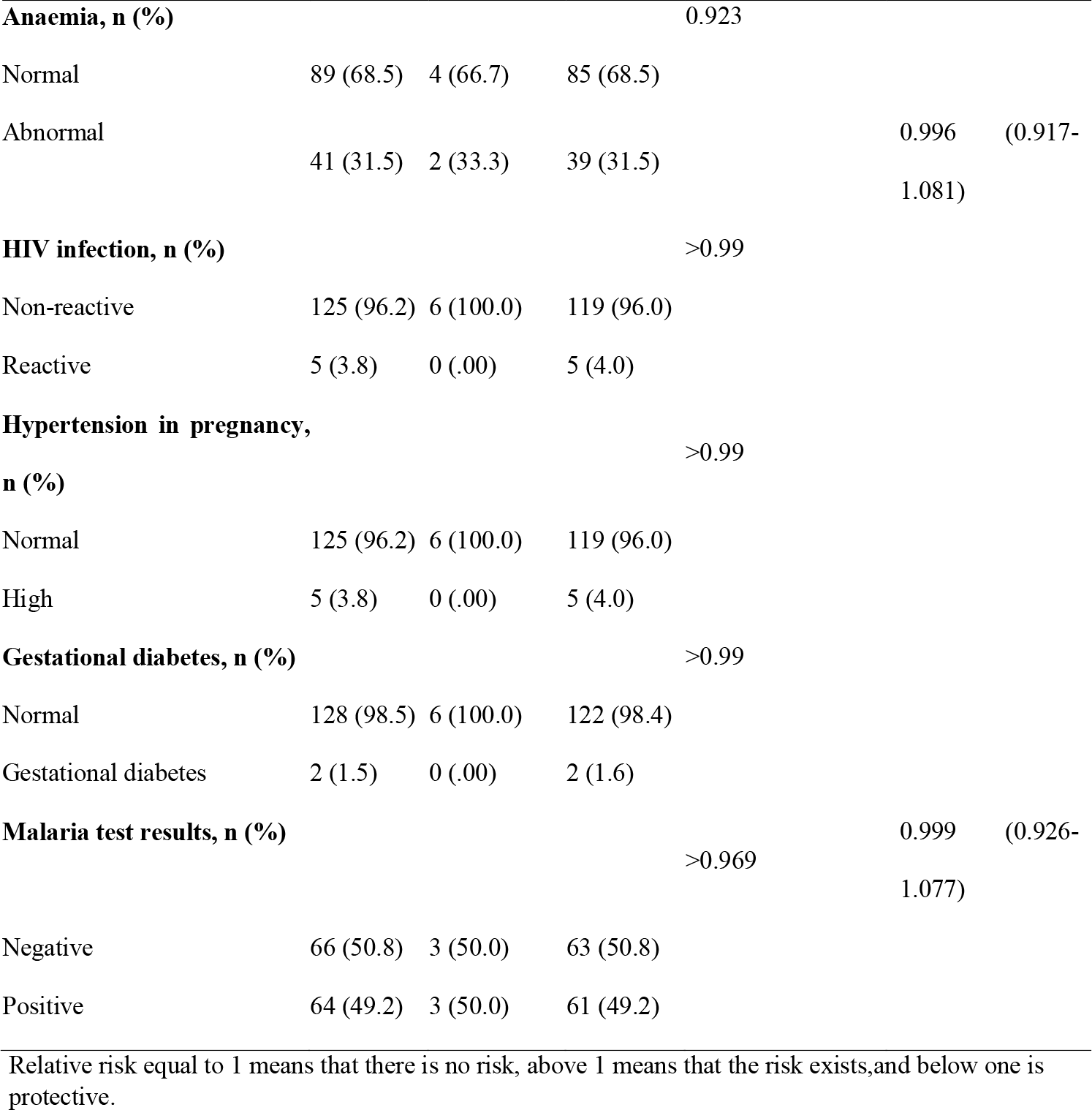
Associated Conditions versus Birth Weight among Pregnant Women. N = 140.

### Associated conditions during pregnancy by malaria results

Malaria test was significantly associated with anemia (p-value <0.001). HIV, hypertension, and gestational diabetes were not associated with malaria test (p-value >0.058). There were 2 (2.9%) positive malaria mothers who had severe malaria. The portion of mothers with HIV reactive results were higher among the positive malaria cohort as compared to the negative cohort with 4 (5.7%) versus 1 (1.4%) respectively. Consequently, there were 5 (7.1%) and 2 (2.9%) of the negative malaria mothers who were pre-eclamptic and had gestational diabetes respectively. None of the positive women had pre-eclampsia nor gestational diabetes. [Table 4]

**Table 4:**
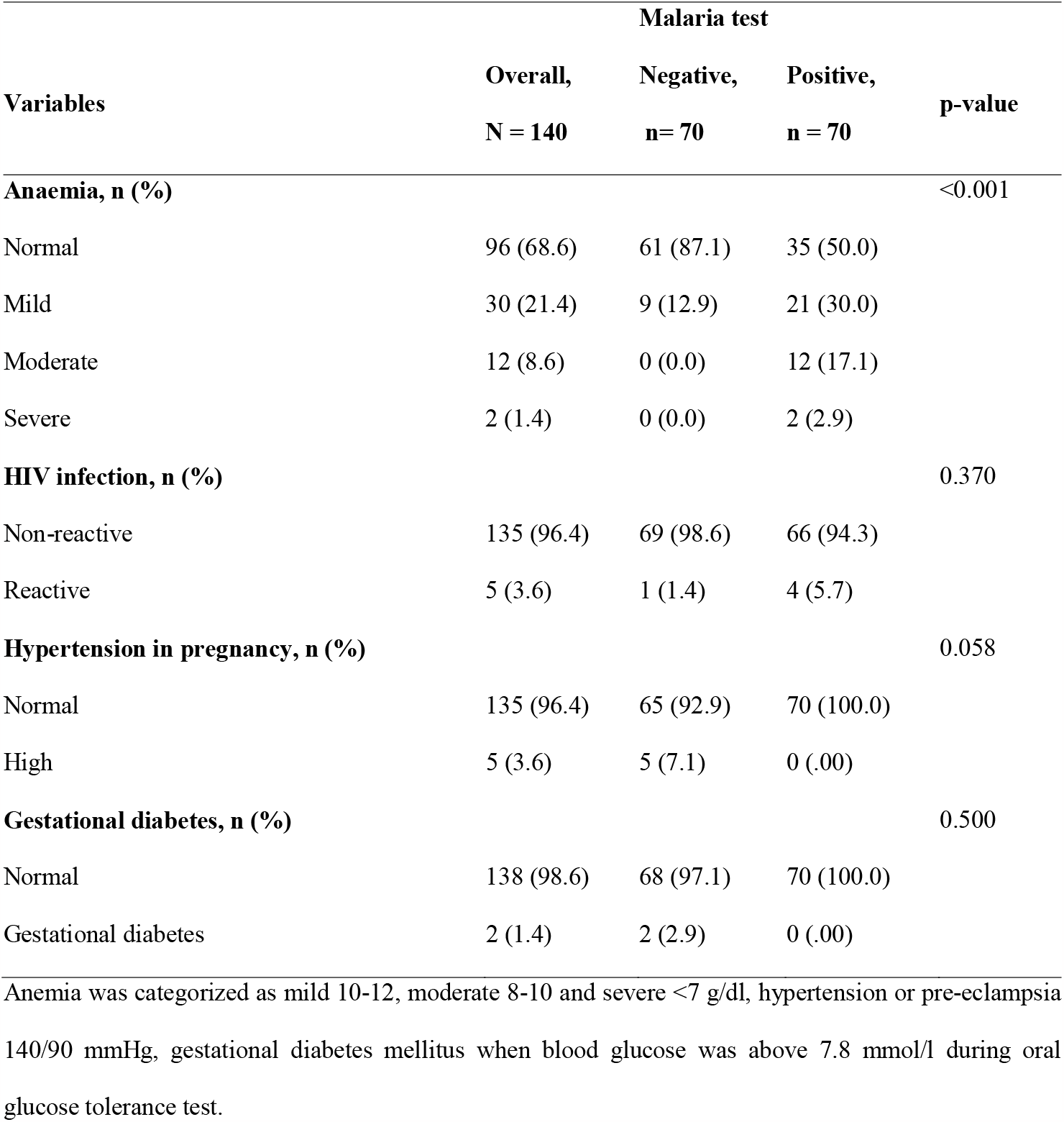
Associated Conditions during Pregnancy by Malaria Results.

## Discussion

Malaria causes low birth weight via a well-known mechanism triggering the placental expression to VAR2CSA, the unique surface antigen responsible of sequestration of plasmodium falciparum evolving in the process of inflammation with vasogenesis, angiogenesis and nutrient transportation dysregulation which affect the fetal development. There is also the severity of malaria effects depending on the level of pre-acquired antimalarial immunity, previous exposure to the infection and the acquired immunity depend on the level of transmission in the residence area. [15], [16], [17]. In the following study, obstetrical causes such as pre-eclampsia, premature rupture of membranes, antepartum hemorrhage, pregnancy age and anemia were associated with low birth weight [18]. We find that in our study malaria test was associated with anemia. In addition, a study found that both use of intermittent preventive treatment and insecticide treated net still are cost-effective methods including barrier protection against contact between mosquitoes and pregnant women blood [19]. In another study the independent or the combined use of both methods was found significantly associated with 20% low birth weight decrease [20]. While another study concluded that early exposure to plasmodium prior to initiating prevention measures was associated with low birth weight [21]. Two or more doses of intermittent preventive treatment were associated with the reduction of low birth weight while one dose of IPT/SP was not associated with the reduction [22].

Notably, studies revealed strong evidence during the last two decades on the decline of malaria concerning its prevalence, clinical cases and disease burden at the community level. This research showed that malaria declined to 40% from 2000-2015 in the Sub-Saharan Africa region. The current result could be attributable majorly to the preventive measures put in place in the healthcare system consisting in the improvement of quality antenatal care through appropriate case management, behaviors and attitudes shaped by social and cultural factors [23]. The fact, quality antenatal care had put emphasize on the early diagnosis of malaria during pregnancy by the mean of microscopy or rapid test, as well as the implementation of the intermittent preventive treatment/sulfadoxine-pyrimethamine and long-lasting treated nets [24], [25].

Studies highlighted that quality interventions were offered to pregnant women during the antenatal clinic, likewise maternal folic acid supplementation in low□and middle□income countries, which was found associated with an increased mean birthweight and decreases in the incidence of low birthweight and small for gestational age. Exposure to more than 4 antenatal visits had decreased incidence of low birth weight [26] [25] [27] [28] [29] [30].

Maternal gravidity and gestational age were key to determine the risk of malaria in the child. It was found statistical association between malaria and low birth weight and other negative consequences more pronounced during the 12 weeks preceding the delivery [31]. As opposed to the previous studies, our findings revealed that the presence of malaria alone or with other illnesses in birth cohorts, did not result in significant negative birth outcomes as similarly corroborated in research carried out in Tanzania and Sudan [32], [33].

Another study concluded that the age, type of place of residence (urban, rural), water sources, marital status, preceding birth interval, sex of newborn, maternal education level, household’s wealth index, access to media, birth order, maternal body mass index, type of cooking fuel, iron supplementation, receipt of antimalarial treatment were predictors associated with low birth weight [34]. Iron and folic acid supplementation was associated with increased birth weight in studies conducted in low-middle income countries [35], [36]. Another study showed that Artemether-lumefantrine was associated with trend towards decreased low birth weight and pregnancy loss [37]. In our study we found that 89% of participants with malaria were treated with Artemether-lumefantrine. Although, current evidence suggesting that the efficacy of antimalarial drugs in preventing low birth weight may decrease with *Plasmodium* resistance, antimalarial medications were used for prevention during pregnancy and showed a significant low birth weight reduction of 27% in the cohort that used the drug when compared with the control group [38]. Malnutrition and malaria share the same geographical area and it contributes to increased disease burden in pregnancy. However, both appear important contributors to low birth weight, and nutrient supplementation during pregnancy appear to be an attractive and feasible intervention to minimize the risk of low birth weight [17]. In a study that analyzed 23 systematic reviews on nutritional interventions during pregnancy, a few factors including provision of vitamin A, low-dose calcium, zinc, and multiple micro-nutrients were associated with reduced risk of low birth weight [39].

Similarly, several earlier micro-nutrient supplementation studies in malarious regions such as Sub-Saharan Africa provided evidence of improved birthweight, increased gestational length and reduced odds of LBW [40]. Evidence suggested that maternal undernutrition is positively associated with low birth weight. In Kenya and Congo Democratic Republic, it was established that the association between malaria infection and reduced fetal growth was greatest among malnourished women. However, in Benin, the effect of malaria infection on fetal growth velocity was greatest among women with low anthropometric status [41], [42]. A Kenyan national survey showed that 2 years after Covid-19 pandemic there was an increase of mother child health service with ANC four visit increasing from 48% to 66% (2009-2022), 88% live births that occurred in the health facility and 89% of delivery that were assisted by a skilled provider. In the meantime, just during the same period there was significant decrease on home delivery estimated at 11% from 34% in the year 2009 [43]. A study carried out in Guinea, malaria immunity for pregnant women was acquired after 6 months of living in the endemic area. Moreover, low immunity induced severe malaria, leading to anemia, abortion and low birth weight [44], and The parasite exposure affected the first baby more than the second baby [22]. Gravidity influenced birth weight reduction, women who were pregnant for the first and second time had significant reduction when compared with multigravida [45], [38]. To support the credence on the above findings, a study conducted in the Sub-Saharan Africa region focused on malaria effect in infants at the delivery highlighted that the first-born child in a context of decreased malaria prevalence was protected against the low birth weight with the use of long-lasting insecticide treated nets [46].

## Conclusion

We observed in this study that birth cohorts did not result in significant low birth weight despite the presence of malaria. This may be due mainly to the concurrent interventions such as antenatal care, public health policy implementation, socioeconomic factors, malaria case management, nutritional status of pregnant women, which have been emphasized on during the last two decades. Therefore, implementing malaria cost-effective strategies in the current context will contribute to good maternal and child health outcomes.

### Limitations

The finding of this research study should be interpreted with caution since the study was carried out at one-site and targeting pregnant women who attended antenatal clinic in the hospital set up. No cohorts of pregnant women and their babies born at home were included.

## Recommendation

Further multiple site longitudinal studies are needed to be carried out in different malaria prone zones including home delivery to determine the effect of each intervention in the context of malaria and low birth weight.

## Data Availability

All data produced in the present study are available upon reasonable request to the authors

## Acknowledgement

We acknowledge the collaboration and efforts of Webuye hospital management team in assisting us during all the process of data collection. Kindly find here the expression of our gratitude.

## Abbreviations

ANC: Antenatal clinic
C/s: caesarian section
CIPD: County Integrated Development Plan
HIV: Human Immunodeficiency Virus
KNBS: Kenya National Bureau of Statistics
Ksh: Kenyan Shilling
RR: Relative Risk
SPSS: Statistical Package for Social Science
WHO: World Health Organization

## Figure legends

Figure 1: Study design

**Figure 1:**
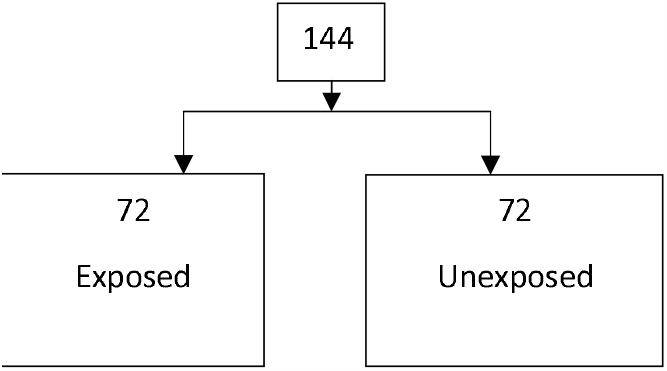
Cohort study with two arms and the expected sample population from the beginning.

Table 1: Socio-demographics characteristics of pregnant women

Table 2: Characteristics of the newborns

Table 3: Associated diseases versus birth weight in pregnancy associated malaria

Table 4: Associated conditions during pregnancy by malaria results

## REFERENCES

1. Sohail, M., Shakeel, S., Kumari, S., Bharti, A., Zahid, F., Anwar, S., Raziuddin, M. (2015). Prevalence of malaria infection and risk factors associated with anaemia among pregnant women in semi urban community of Hazaribag, Jharkhand, India. BioMed research international.

2. World Health Organization. (2023). WHO guidelines for malaria, 14 March 2023 (No. WHO/UCN/GMP/2023.01). World Health Organization.

3. Kenya Malaria Indicator Survey. (2019-2023). National Malaria Control Programme, Ministry of Health, Kenya National Bureau of Statistics, ICF International.

4. WHO (2019). The “World malaria report 2019” at a glance. World Health Organization, Geneva, Switzerland.

5. WHO (2014). Policy brief for the implementation of intermittent preventive treatment of malaria in pregnancy using sulfadoxine-pyrimethamine (IPTp-SP). April 2013 (revised January 2014).

6. Imboumy-Limoukou, R. K., Maghendji-Nzondo, S., Sir-Ondo-Enguier, P. N., De Carvalho, J. N., Tsafack-Tegomo, N. P., Buekens, J., Lekana-Douki, J. B. (2020). Malaria in children and women of childbearing age: infection prevalence, knowledge and use of malaria prevention tools in the province of Nyanga, Gabon. Malaria journal, 19(1), 1–8.

7. Alonso, P. L., Noor, A., Santelli, A. C., Ghani, A., de Quadros, C., Karema, C., Reeder, J. (2015). Global technical strategy for malaria 2016–2030.

8. Zhou, X. N. (2021). China declared malaria-free: a milestone in the world malaria eradication and Chinese public health.

9. National Malaria Control Programme-NMCP/Kenya, Kenya National Bureau of Statistics-KNBS, and ICF International.2016. Kenya Malaria Indicator Survey 2015. Nairobi, Kenya.

10. Gueneuc, A., Deloron, P., Bertin, G. I. (2017). Usefulness of a biomarker to identify placental dysfunction in the context of malaria. Malaria journal, 16, 1–7.

11. Wekesa, A, Mulambalah, C, Mulama, D, Elizabeth, O. (2019). Prevalence and risk analysis among pregnant women in Bungoma County. Medicine science, International medical journal.

12. DHIS2, District Health Information Systems. 2019.

13. Bungoma County Integrated Development Plan 2018-2022

14. Nyamu, G. W., Kihara, J. H., Oyugi, E. O., Omballa, V., El-Busaidy, H., Jeza, V. T. (2020). Prevalence and risk factors associated with asymptomatic Plasmodium falciparum infection and anemia among pregnant women at the first antenatal care visit: A hospital based cross-sectional study in Kwale County, Kenya. PloS one, 15(10), e0239578.

15. Guyatt, H. L., Snow, R. W. (2004). Impact of malaria during pregnancy on low birth weight in sub-Saharan Africa. Clinical microbiology reviews, 17(4), 760–769.

16. Chua, C. L., Hasang, W., Rogerson, S. J., Teo, A. (2021). Poor birth outcomes in malaria in pregnancy: recent insights into mechanisms and prevention approaches. Frontiers in immunology, 12, 621382.

17. Ismail, M. R., J. Ordi, C. Menendez, P. J. Ventura, J. J. Aponte, E. Kahigwa, R. Hirt, A. Cardesa, and A. P. L. Alonso. (2000). Placental pathology in malaria: a histological, immunohistochemical, and quantitative study. Hum. Pathol.31:85–93). (Mohammed et al., 2013.

18. Alebel, A., Wagnew, F., Tesema, C., Gebrie, A., Ketema, D. B., Asmare, G., Kibret, G. D. (2019). Factors associated with low birth weight at Debre Markos Referral Hospital, Northwest Ethiopia: a hospital based cross-sectional study. BMC research notes, 12, 1–6.

19. Manu, G., Boamah-Kaali, E. A., Febir, L. G., Ayipah, E., Owusu-Agyei, S., Asante, K. P. (2017). Low utilization of insecticide-treated bed net among pregnant women in the middle belt of Ghana. Malaria research and treatment, 2017.

20. Nkoka, O., Chuang, T. W., Chen, Y. H. (2020). Effects of malaria interventions during pregnancy on low birth weight in Malawi. American Journal of Preventive Medicine, 59(6), 904–913.

21. Schmiegelow, C., Matondo, S., Minja, D. T., Resende, M., Pehrson, C., Nielsen, B. B., Theander, T. G. (2017). Plasmodium falciparum infection early in pregnancy has profound consequences for fetal growth. The Journal of infectious diseases, 216(12), 1601–1610.

22. Manyando, C., Njunju, E. M., Mwakazanga, D., Chongwe, G., Mkandawire, R., Champo, D., Van Geertruyden, J. P. (2014). Safety of daily co-trimoxazole in pregnancy in an area of changing malaria epidemiology: a phase 3b randomized controlled clinical trial. PloS one, 9(5), e96017.

23. Bhatt, S., Weiss, D. J., Cameron, E., Bisanzio, D., Mappin, B., Dalrymple, U., Gething, P. W. (2015). The effect of malaria control on Plasmodium falciparum in Africa between 2000 and 2015. Nature, 526(7572), 207–211.

24. Uwimana, G., Elhoumed, M., Gebremedhin, M. A., Azalati, M. M., Nan, L., Zeng, L. (2023). Association between quality antenatal care and low birth weight in Rwanda: a cross-sectional study design using the Rwanda demographic and health surveys data. BMC Health Services Research, 23(1), 1–10.

25. Calliope, S. A., Yorifuji, T., Wada, T., Mukakarake, M. G., Mutesa, L., Yamamoto, T. (2020). Antenatal care visits and adverse pregnancy outcomes at a hospital in rural western province, Rwanda. Acta Medica Okayama, 74(6), 495–503.

26. Jonker, H., Capelle, N., Lanes, A., Wen, S. W., Walker, M., Corsi, D. J. (2020). Maternal folic acid supplementation and infant birthweight in low□and middle□income countries: A systematic review. Maternal & child nutrition, 16(1), e12895.

27. Kassar, S. B., Melo, A. M., Coutinho, S. B., Lima, M. C., Lira, P. I. (2013). Determinants of neonatal death with emphasis on health care during pregnancy, childbirth and reproductive history. Jornal de pediatria, 89(3), 269–277.

28. Khatun, S., Rahman, M. (2008). Socio-economic determinants of low birth weight in Bangladesh: a multivariate approach. Bangladesh Medical Research Council Bulletin, 34(3), 81–86.

29. Khan, N., Jamal, M. (2003). Maternal risk factors associated with low birth weight. Journal of the College of Physicians and Surgeons--pakistan: JCPSP, 13(1), 25–28.

30. Kananura, R. M., Wamala, R., Ekirapa-Kiracho, E., Tetui, M., Kiwanuka, S. N., Waiswa, P., Atuhaire, L. K. (2017). A structural equation analysis on the relationship between maternal health services utilization and newborn health outcomes: a cross-sectional study in Eastern Uganda. BMC pregnancy and childbirth, 17(1), 1–12.

31. Beaudrap, P., Turyakira, E., Nabasumba, C., Tumwebaze, B., Piola, P., Boum Ii, Y., Gready, R. (2016). Timing of malaria in pregnancy and impact on infant growth and morbidity: a cohort study in Uganda. Malaria journal, 15, 92. 10.1186/s12936-016-1135-7.

32. Kalinjuma, A. V., Darling, A. M., Mugusi, F. M., Abioye, A. I., Okumu, F. O., Aboud, S., Fawzi, W. W. (2020). Factors associated with sub-microscopic placental malaria and its association with adverse pregnancy outcomes among HIV-negative women in Dar es Salaam, Tanzania: a cohort study. BMC Infectious Diseases, 20(1), 1–13.

33. Mohammed, A. H., Salih, M. M., Elhassan, E. M., Mohmmed, A. A., Elzaki, S. E., El-Sayed, B. B., & Adam, I. (2013). Submicroscopic Plasmodium falciparum malaria and low birth weight in an area of unstable malaria transmission in Central Sudan. Malaria journal, 12, 1–6.

34. Babitsch, B., Gohl, D., Von Lengerke, T. (2012). Re-revisiting Andersen’s Behavioral Model of Health Services Use: a systematic review of studies from 1998–2011. GMS Psycho-Social-Medicine, 9.

35. Caniglia, E. C., Zash, R., Swanson, S. A., Smith, E., Sudfeld, C., Finkelstein, J. L., Shapiro, R. L. (2022). Iron, folic acid, and multiple micronutrient supplementation strategies during pregnancy and adverse birth outcomes in Botswana. The Lancet Global Health, 10(6), e850–e861.

36. Kamau, M. W., Mirie, W., Kimani, S. (2018). Compliance with Iron and folic acid supplementation (IFAS) and associated factors among pregnant women: results from a cross-sectional study in Kiambu County, Kenya. BMC public health, 18, 1–10.

37. Muehlenbachs, A., Fried, M., McGready, R., Harrington, W. E., Mutabingwa, T. K., Nosten, F., Duffy, P. E. (2010). A novel histological grading scheme for placental malaria applied in areas of high and low malaria transmission. The Journal of infectious diseases, 202(10), 1608–1616.

38. Muanda, F. T., Chaabane, S., Boukhris, T., Santos, F., Sheehy, O., Perreault, S., Bérard, A. (2015). Antimalarial drugs for preventing malaria during pregnancy and the risk of low birth weight: a systematic review and meta-analysis of randomized and quasi-randomized trials. BMC medicine, 13(1), 1–14).

39. da Silva Lopes, K., Ota, E., Shakya, P., Dagvadorj, A., Balogun, O. O., Peña-Rosas, J. P., Mori, R. (2017). Effects of nutrition interventions during pregnancy on low birth weight: an overview of systematic reviews. BMJ global health, 2(3), e000389.

40. Unger, H. W., Ashorn, P., Cates, J. E., Dewey, K. G., Rogerson, S. J. (2016). Undernutrition and malaria in pregnancy–a dangerous dyad? BMC medicine, 14, 1–9.

41. Cates, J.E., Unger, H.W., Briand, V., Fievet, N., Valea, I., Tinto, H., Rogerson, S. (2017). Malaria, malnutrition, and birthweight: a meta-analysis using individual participant data. PLoS medicine, 14(8), e1002373.

42. Landis, S. H., Lokomba, V., Ananth, C. V., Atibu, J., Ryder, R. W., Hartmann, K. E., Meshnick, S. R. (2009). Impact of maternal malaria and under-nutrition on intrauterine growth restriction: a prospective ultrasound study in Democratic Republic of Congo. Epidemiology & Infection, 137(2), 294–304.

43. KNBS and ICF. 2023. Kenya Demographic and Health Survey 2022: Volume 1. Nairobi, Kenya, and Rockville, Maryland, USA: KNBS and ICF.

44. Touré, A A., Abdoulaye, D., Abdourahamane, D., Gaspard, L., Abdourahim, C., Sidikiba, S., Abdoul, H B. (2019). Malaria associated factors among pregnant women in Guinea. Journal of tropical medicine, 2019.

45. Kayentao, K., Garner, P., van Eijk, A. M., Naidoo, I., Roper, C., Mulokozi, A., Kuile, F. O. (2013). Intermittent preventive therapy for malaria during pregnancy using 2 versus 3 or more doses of sulfadoxine-pyrimethamine and risk of low birth weight in Africa: systematic review and meta-analysis. Jama, 309(6), 594–604.

46. Heng, S., O’Meara, W. P., Simmons, R. A., Small, D. S. (2021). Relationship between changing malaria burden and low birth weight in sub-Saharan Africa: A difference-in-differences study via a pair-of-pairs approach. Elife, 10, e65133.

